# Using large language models to understand the public discourse towards vaccination in Brazil between January 2013 and December 2019

**DOI:** 10.1101/2025.02.24.25322766

**Authors:** Laura Espinosa, Marcel Salathé

## Abstract

**Background:** Vaccination against infectious diseases prevents diseases, saves lives, and reduces healthcare costs. However, trust, accessibility, and public perceptions influence vaccine acceptance or refusal. Risk communication and community engagement (RCCE) is key in addressing inaccurate claims and misconceptions, and improving vaccine coverage. This study examines how public stance towards vaccination evolved in Brazil from 2013 to 2019 using Twitter data.

**Methods:** We collected tweets in Portuguese containing vaccine-related keywords from January 2013 to December 2019 using Crowdbreaks. We used Python and GPT-4 with few-shot classification to extract the stance towards vaccination. We performed a descriptive analysis, including time and geographical components, of the stance towards vaccination. Data on measles-mumps-rubella (MMR) vaccination coverage and confirmed measles cases were also analyzed. Pairwise trend comparisons explored relationships between stance, vaccination coverage, and measles incidence.

**Findings:** We collected 2,197,090 tweets with 1,703,009 classified by stance. Most tweets were neutral (47%), but negative stance increased over time peaking in 2019. This decline in positive stance coincided with the resurgence of measles in 2018-2019. Some states like Rio de Janeiro and Rio Grande do Sul had a consistently lower Twitter vaccine stance (TVS) index. While no clear pattern emerged from trend comparisons, increased measles cases were often followed by a decreased TVS index.

**Interpretation:** Public stance towards vaccination showed a noticeable geographical and temporal variation. The decline in TVS index, suggesting a possible relation with the decline in vaccination coverage, highlights the need for sustained and continuous public health efforts. Monitoring public stance in real-time can support RCCE strategies to improve vaccine confidence and uptake. Future research should integrate offline data and examine how epidemics influence public health stance.

**Research into context:** 

**Evidence before this study:** Understanding the perception of vaccines by the population is one of the key determinants for ensuring appropriate vaccination coverage and successful prevention and control of vaccine-preventable diseases. Surveys used to be the traditional method to collect this information; however, public social media data allow for a more scalable approach to collect this information. Previous studies have used conceptual models, such as the “3Cs” (Complacency, Convenience, and Confidence), to understand vaccine hesitancy. Additionally, large-scale social media analyses have provided insights into vaccine-related discussions, though traditional surveys often suffer from biases and delayed reporting.

**Added value of this study:** This study leverages large-scale social media data and large language models (LLMs), specifically GPT-4, to classify public stance toward vaccination over seven years in Brazil. It shows the feasibility of using real-time social media analysis to track vaccine-related discourse, revealing trends in vaccine stance, and providing actionable insights. The study provides a detailed understanding of vaccine stance across different geographical and temporal components, identifying regions where negative stance is more pronounced.

**Implications of all the available evidence:** The findings highlight the importance of continuously monitoring vaccine stance to support successful and targeted public health interventions and RCCE strategies. While this study focused on a specific social media platform, time, and place, this approach can be easily applied to other data sources, contexts, and time periods.

## 1. Introduction

Vaccination is one of the greatest achievements in public health of the 20th century. Vaccination against infectious diseases not only saves lives, improves individual and population health, but also saves billions of dollars annually. Despite all this, there are persistent issues related to inadequate, delayed and unstable vaccination uptake.^1^

Understanding the vaccination determinants, including enablers and drivers, is key to improve vaccination coverage to reach sufficient levels for the best health outcomes. Depending on how transmissible a disease is, the vaccination coverage required to provide herd immunity or disease eradication will change. For example, measles, as one of the most contagious vaccine-preventable diseases, requires more than 90% of vaccination coverage to reach herd immunity; and rubella is estimated to require more than 80% of vaccination coverage as a protection threshold.^2^

In October 2014, World Health Organization (WHO) Strategic Advisory Group of Experts on Immunization (SAGE) published a report of the SAGE Working Group on Vaccine Hesitancy outlying several factors that can influence vaccine acceptance or refusal.^3^ The working group assessed several conceptual models on vaccine hesitancy determinants, focusing on their complexity and global applicability.^4^ The working group concluded that vaccine hesitancy is not driven by a simple set of individual factors; the Complacency, Convenience and Confidence model (‘3Cs’) was intuitive and a more comprehensive matrix capturing the contextual, individual, group, and vaccine/vaccination-specific influences was developed. Some of the determinants influencing vaccine acceptance or refusal are related to the trust and self-perception by the population to the effectiveness and safety of vaccines and the system that delivers these vaccines, including the health system, health professionals, and policy makers deciding on which vaccines to administer to the population.

Effective public health communication and Risk Communication and Community Engagement (RCCE) are crucial for enhancing vaccination uptake. RCCE principles of transparency, trust-building, cultural sensitivity, two-way communication, and community participation help create an environment where people can access accurate information and make informed decisions about vaccination.^5,6^

Understanding public discourse toward vaccination is essential for tailoring risk communication strategies to enhance and maintain vaccination uptake. For example, social media data was analyzed to assess public stance toward vaccination in relation to measles in Europe. This non-invasive approach can help identifying regions with varying levels of vaccine acceptance, enabling community-specific initiatives to provide accurate information and support informed vaccination decisions.^7,8^ This approach complements traditional surveys, which are often limited by biases (e.g., increased agreement rates) and delayed reporting.^9^

Being able to have near-real-time data on public discourse towards vaccination is critical for incorporating it into RCCE strategies ensuring a more responsive and impactful approach to public health interventions and challenges.

In 2016, Brazil achieved a significant public health milestone when it was declared measles-free following the WHO criteria: absence of endemic measles virus transmission for at least 36 consecutive months, verified standard surveillance and genotyping evidence supporting the interruption of the transmission.^10,11^ This milestone was part of a broader success, as the region of the Americas became the first WHO region to be certified as measles-free.^12,13^ However, in 2018, measles re-emerged in Brazil, which led to the loss of Brazil’s measles-free status in 2019.

During the same period, Brazil faced an epidemic of yellow fever, which started at the end of 2016 and peaked in 2018.^14^ This further strained the public health infrastructure and resources in Brazil, and underscored the need for timely immunisation campaigns accompanied with targeted public health communication. This study aims at understanding the public stance towards vaccination and vaccines of Twitter users in Brazil between 2013 and 2019 and assessing the discourse behind each stance and time period.

## 2. Materials and methods

### 2.1. Twitter data collection

Crowdbreaks (ref) was used for Twitter data collection.

The following keywords in Portuguese were used to collect tweets from January 2013 to December 2019 through the Twitter API version 1.1 endpoint POST status/filter (ref): ‘vacina’, ‘vacinação’, ‘vacinado’, ‘vacinar’, and ‘vacinal’.

### 2.2. Public stance towards vaccination expressed in tweets text

#### 2.1.1. Extracting the public stance towards vaccination expressed in tweets text

We used Python 3.9.18 and GPT-4 with few-shot classification due to its good performance in this type of use cases shown in a previous study to classify the tweets collected according to the public stance towards vaccination.^15^

#### 2.2.2. Descriptive analysis of public stance towards vaccination expressed in tweets text

We calculated the Twitter vaccine stance (TVS) index as follows:

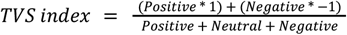

We performed a descriptive analysis, including time and geographical components, of the stance towards vaccination using R 4.1.3 and RStudio version 2023.06.1. The code and list of R packages used are available in the repository ‘brazil-vaccine-uptake’.

Furthermore, we used GPT-4-o-mini to summarise a maximum of 2,450 randomly selected tweets per month, year and stance based on the distribution per week and Brazilian state. This limitation in the number of tweets was based on the limit number of input tokens that GPT-4-o-mini had for the prompt.

### 2.3. Measles-mumps-rubella (MMR) vaccination coverage of the first dose and confirmed measles cases

The MMR vaccination coverage of the first dose was collected from Boccolini *et al*., 2023 and it was calculated with the number of final vaccine doses and estimated target population from official sources, excluding values bigger than 150% to avoid implausible outliers.^16^ The number of confirmed measles cases per year and Brazilian state were collected from the epidemiological reports of the Brazilian Ministry of Health and were standardised based on the population per state and year.^17^ We performed a descriptive analysis, including time and geographical components, of the confirmed measles cases and MMR vaccination coverage of the first dose using R 4.1.3 and RStudio version 2023.06.1.

### 2.4. Comparison of trends

We calculated the qualitative (decreased, increased or equal) and quantitative (percentage difference) trend per year and Brazilian state for the public stance, MMR vaccination coverage of the first dose and confirmed measles cases. Moreover, we calculated the pairwise comparison of the qualitative trends of these three indicators per year and state with a lag of zero, one and two years for the second value in the pairwise comparison. Then, we aggregated these pairwise comparisons and calculated the percentage per pairwise comparison and lag of the different possible combinations. We excluded the year 2013 for public stance since there was no data for 2012 to calculate the trend, as well as the equal trends for confirmed measles cases which are due to having no cases for more than one year in a row.

### 2.5. Ethics

This work has been approved by the EPFL Human Research Ethics Committee (HREC No. 012.2018 / 05.03.2018).

Crowdbreaks anonymises all tweets’ text changing the mention or reference to usernames to “@username” and the URLs to “url”, and complies with the Twitter terms of service.

## 3. Results

### 3.1. Twitter data collection

A total of 2,197,090 tweets in Portuguese with information on user geolocation from Brazil containing the keywords were collected between 1 January 2013 and 31 December 2019. From these, 1,750,294 tweets had information on location at least at the state level which were used for the geographical maps.

### 3.2. Public stance towards vaccination expressed in tweets text

#### 3.2.1. Extracting the public stance towards vaccination expressed in tweets text

A Portuguese few-shot classification prompt was used with GPT-4 to extract the public stance towards vaccination expressed in tweets text. From the total number of tweets collected, 1,703,009 tweets were properly classified by GPT-4 (e.g., for some tweets, GPT-4 did not provide a stance towards vaccination) and, from these, 1,312,792 tweets had information on location at least at the state level.

#### 3.2.2. Descriptive analysis of public stance towards vaccination expressed in tweets text

Overall, public stance was found 47% neutral (ranging from 39·9% in 2019 to 54·5% in 2016), 26% negative (ranging from 14·1% in 2015 to 32·7% in 2019) and 28% positive (ranging from 25·1% in 2018 to 32·9% in 2015) (Table 1).

**Table 1.** Summary of total number of tweets per year and class of public stance towards vaccination.

| Characteristic | 2013, N =<br>109,228<sup>†</sup> | 2014, N =<br>127,478<sup>†</sup> | 2015, N =<br>107,840<sup>†</sup> | 2016, N =<br>142,485<sup>†</sup> | 2017, N =<br>180,358<sup>†</sup> | 2018, N =<br>371,110<sup>†</sup> | 2019, N =<br>664,510<sup>†</sup> |
| --- | --- | --- | --- | --- | --- | --- | --- |
| Stance |  |  |  |  |  |  |  |
| negative | 20,795 (19%) | 26,730 (21%) | 15,152 (14%) | 24,539 (17%) | 38,129 (21%) | 91,807 (25%) | 217,413 (33%) |
| neutral | 52,987 (49%) | 63,331 (50%) | 57,242 (53%) | 77,684 (55%) | 95,831 (53%) | 186,255 (50%) | 265,019 (40%) |
| positive | 35,446 (32%) | 37,417 (29%) | 35,446 (33%) | 40,262 (28%) | 46,398 (26%) | 93,048 (25%) | 182,078 (27%) |
| <sup>†</sup> n (%) |  |  |  |  |  |  |  |

The seven-day moving average of the TVS index showed that at national level the majority of the 2,513 days were above zero ranging from 56·3% in 2019 to 98·1% in 2015 (Fig 1). At state level, Minas Gerais and São Paolo had the highest number of tweets and a similar TVS index values as national level and no tweets were collected for Rio Grande Do Norte. Moreover, the seven-day moving average of the TVS index showed a similar trend at state level than national, except for Rio de Janeiro where 51·3% of the 2,513 days were below zero.

**Figure 1.**
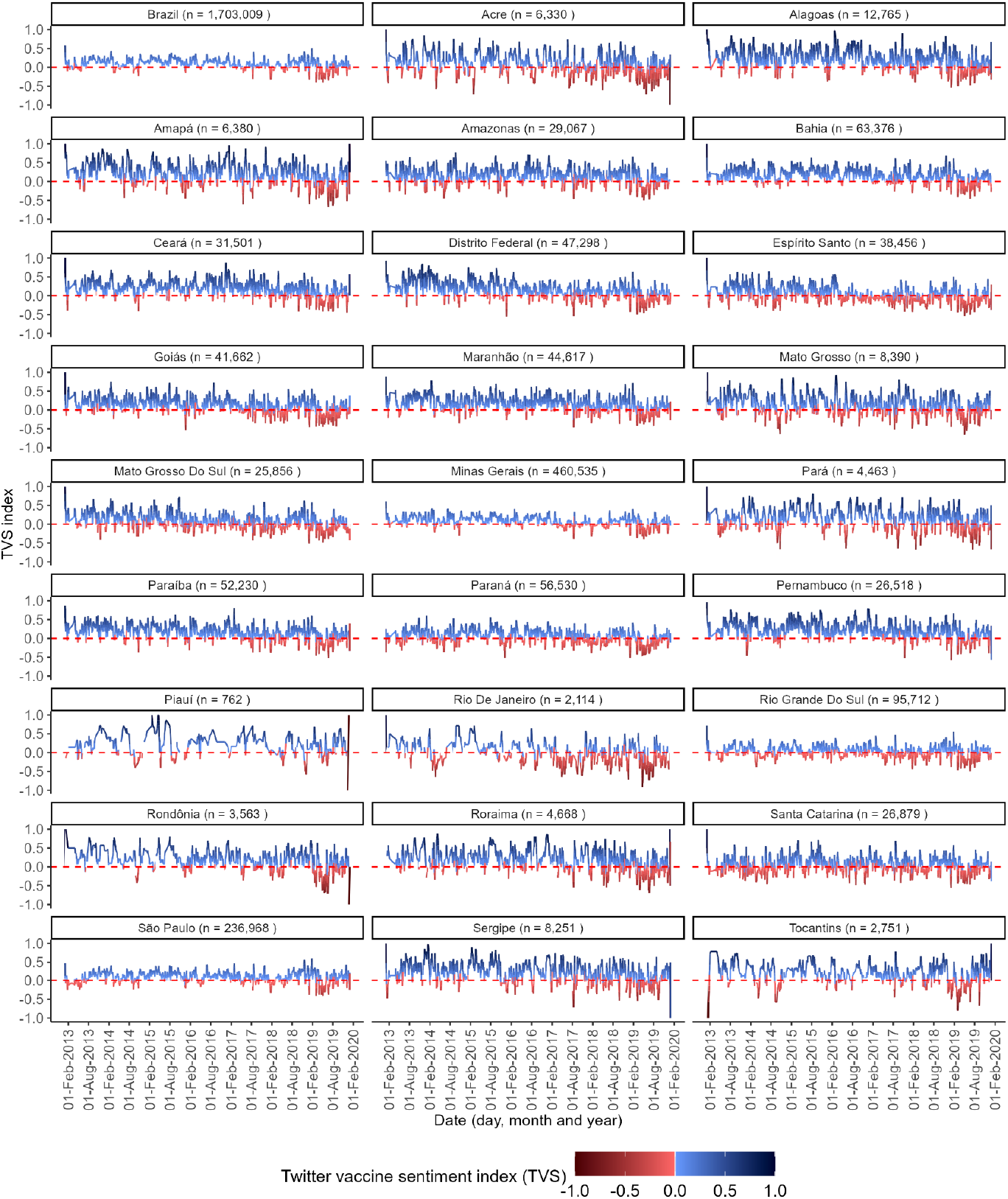
Seven-day moving average of TVS index in Brazil and per Brazilian state, January 2013 to December 2019.

Figure 2 shows the distribution of TVS index per year and Brazilian state. In terms of distribution of TVS index per Brazilian state, Rio Grande do Sul presents a negative TVS index in all years except in 2015, Santa Catarina and Rio de Janeiro had negative TVS index in three additional years apart from 2019, and Minas Gerais and Sao Paolo had negative TVS index in two additional years apart from 2019. In terms of distribution of TVS per year, all states had a positive TVS index in 2015 and, from 2016 and especially in 2019, there are more states with negative TVS index (18 of the 25 states with data) and also the only year with a negative median (−0·04) and the lowest maximum (0·13). The TVS index ranged from -0·19 in 2017 to 0·43 in 2015 and from -0·19 in Rio de Janeiro to 0·43 in Sergipe.

**Figure 2.**
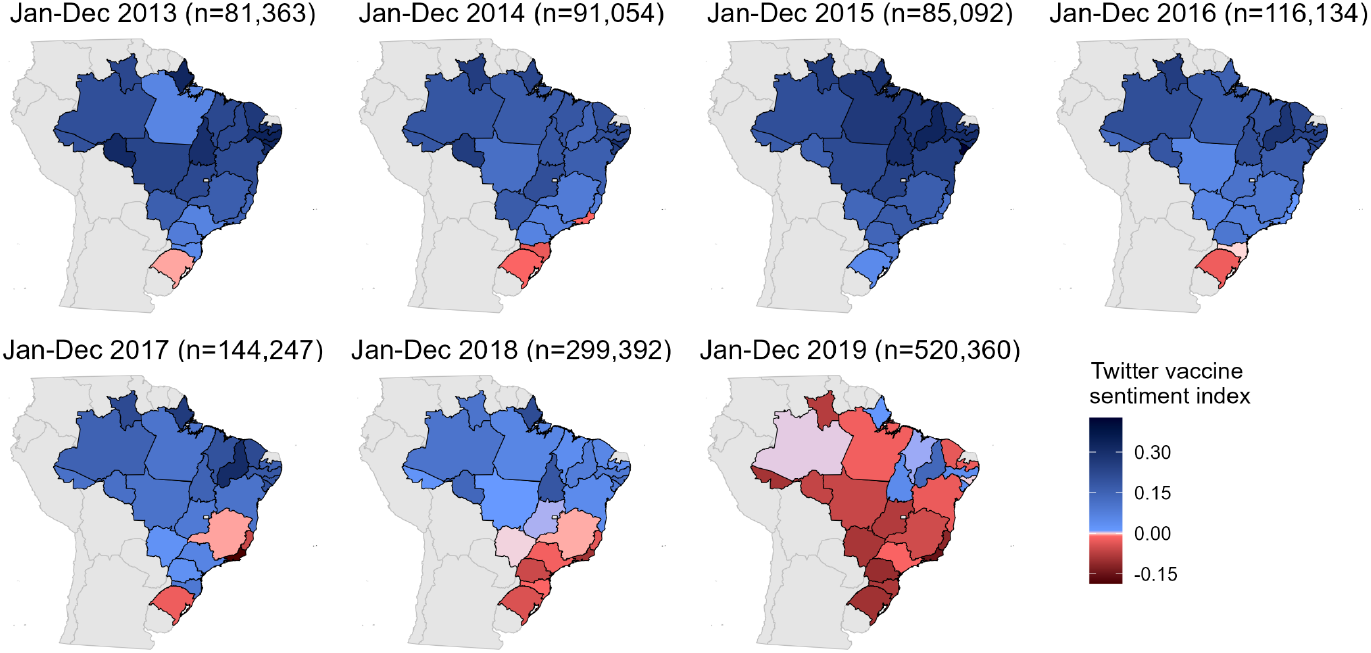
TVS index per year and Brazilian state, January 2013 to December 2019 (N = 1,337,642).

The summaries of the texts from selected tweets per stance, year, and month revealed recurrent themes and key narratives (Supplementary material). In the tweets with a positive stance towards vaccination, tweets frequently appraised the effectiveness of vaccines and importance of vaccination campaigns for disease prevention and control, especially during an epidemic such as the yellow fever epidemic in 2018. In contrast, the tweets with a negative stance towards vaccination often referred to incorrect information about vaccine safety, distrust of the health and national authorities, and concerns about the ongoing measles outbreak and distrust in the vaccine as a mechanism to end it. Lastly, tweets with a neutral stance towards vaccination focused on factual updates or announcements about vaccination schedules, logistical information about vaccination campaigns or general public health messages.

### 3.3. Descriptive analysis of MMR vaccination coverage in the first dose and confirmed measles cases

The MMR vaccination coverage for the first dose ranged from 68·0% to 145·7% (Fig. 3). The percentage above 100%, according to the authors of the dataset, can be likely due to errors in the numerator (i.e., number of vaccine doses) or denominator (i.e., estimation of the target population), mid-year modifications in the target population or the inclusion of children from other cities in the numerator. Only seven states (Alagoas, Ceara, Mato Grosso do Sul, Minas Gerais, Pernambuco, Rio de Janeiro, and Rondonia) reached the WHO target of 95% MMR vaccination coverage for the first dose in all years, whereas 14 states reached that goal only in two or three years of the study period. In 2014, all states reached the 95% goal, whereas 2018 and 2019 had only nine or ten states, respectively, reaching the 95% goal.

**Figure 3.**
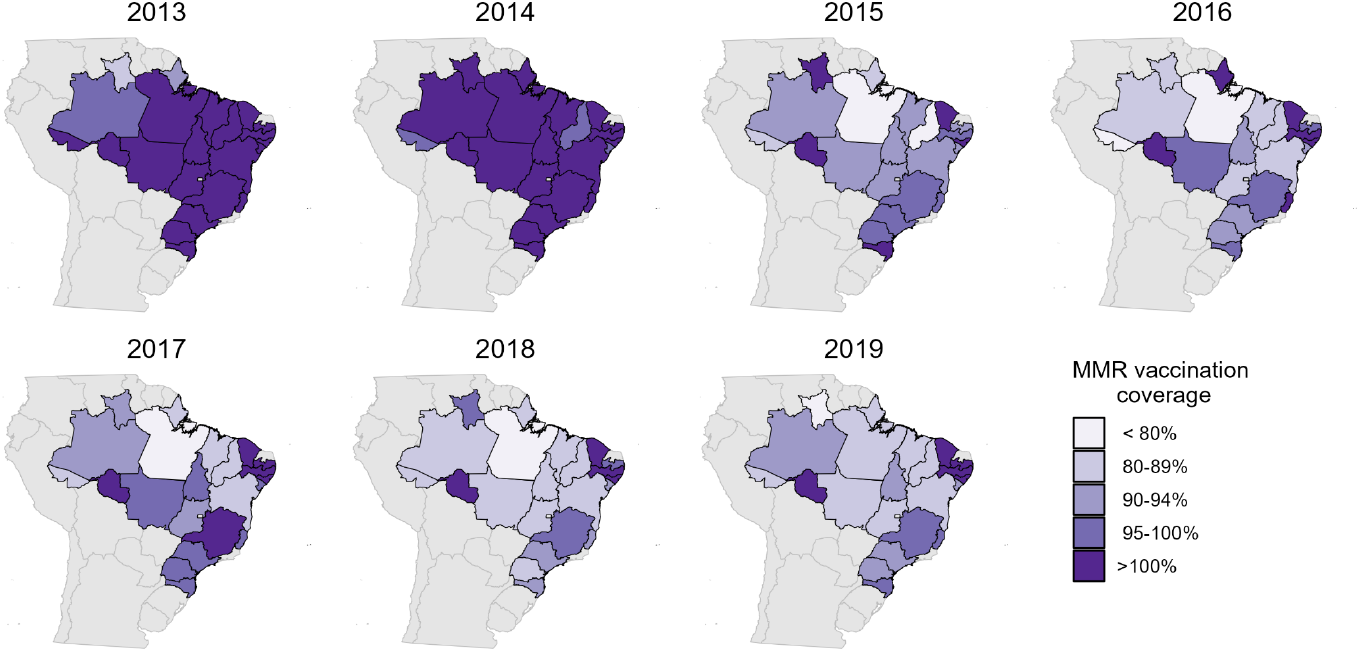
MMR vaccination coverage for the first dose per year and Brazilian state, 2013 to 2019.

The number of confirmed measles cases per 100,000 population per year and state ranged from 0 to 215·4, with an interquartile range of 77 cases per 100,000 population. Figure 4 shows the geographical and yearly distribution of confirmed measles cases based on the four quartiles. Until 2018, there were very few cases and in 2018 and 2019, the number of states reporting cases and number of cases per state increased dramatically.

**Figure 4.**
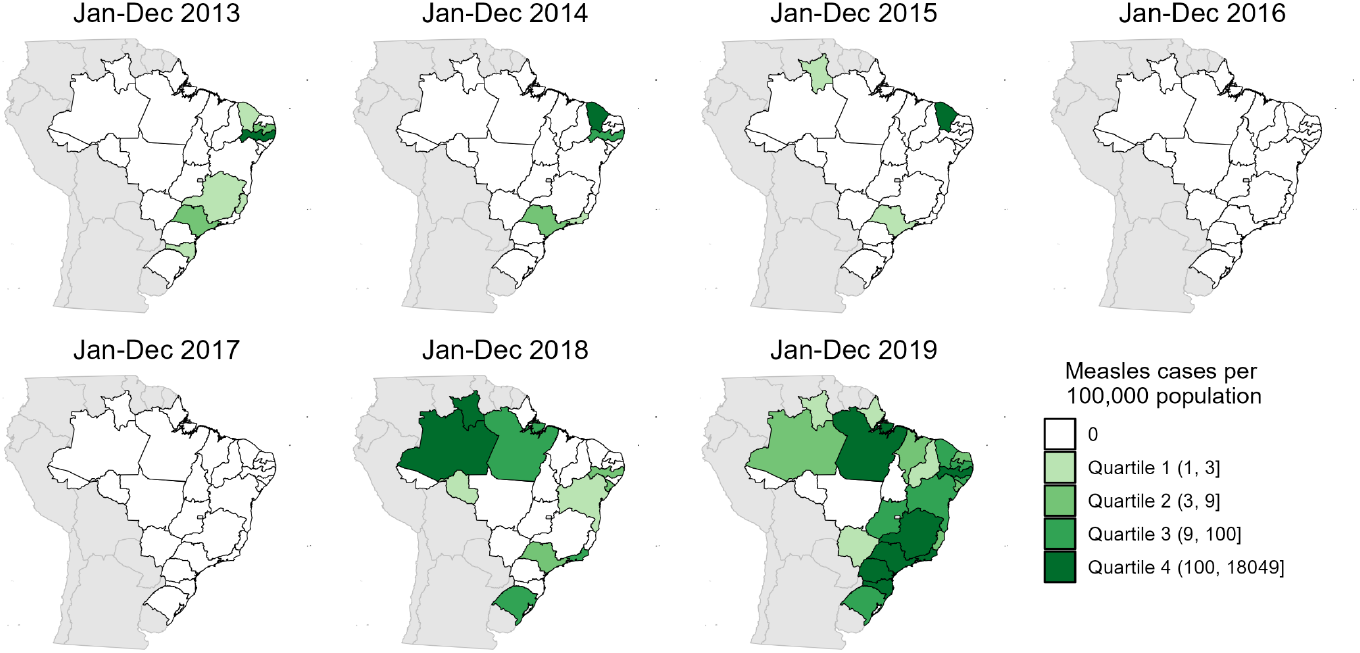
Number of measles cases per year and Brazilian state, 2013 to 2019.

### 3.4. Analysis of the trend comparison

Figure 5 shows the trends of the three indicators included in this study per year and Brazilian state: TVS index, MMR coverage in the first dose and confirmed cases of measles. Panel A illustrates the growing proportion of states showing a decrease in TVS index, particularly between 2017 and 2019, while states with increased TVS index were more or less consistent between 2017 and 2019. Panel B shows an intermittent pattern of coverage across states, with most states experiencing increased coverage in 2014, 2017, and 2019, while other years exhibit decreased coverage. Panel C highlights the resurgence of measles cases in 2018 when 11 states showed an increased trend. The widespread increased trend of measles cases in 2018 is preceded by some years of decreased MMR coverage, including in 2018, and decreased TVS index.

**Figure 5.**
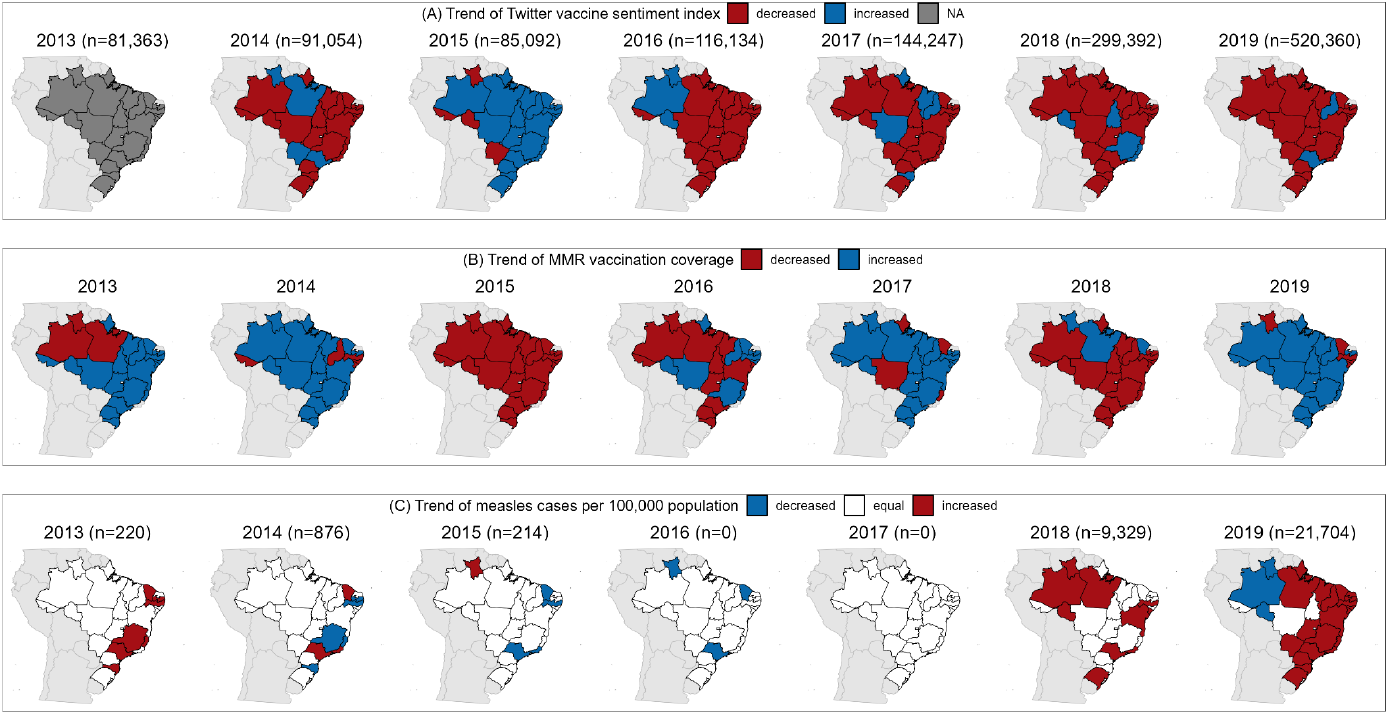
Trends of (A) TVS index, (B) MMR coverage and (C) confirmed cases of measles, 2013-2019.

There were 3,294 values in total for the pairwise comparison per year and state. When excluding the not applicable values for not being able to calculate the trend of TVS in 2013, there were 2,600 values. Furthermore, when excluding the equal trends for confirmed measles cases, there were a total of 1,346 values. Overall, there were no clear patterns on the predominant qualitative trends combination for the pairwise comparisons (Table 2). Nonetheless, the pairwise comparison of confirmed measles cases trends with a lag of zero, one and two years of the TVS index trends showed a predominance of increased confirmed measles cases followed by decreased TVS trends in the same year or one or two years after (58·8%, 66·7% and 64·9%, respectively).

**Table 2.**
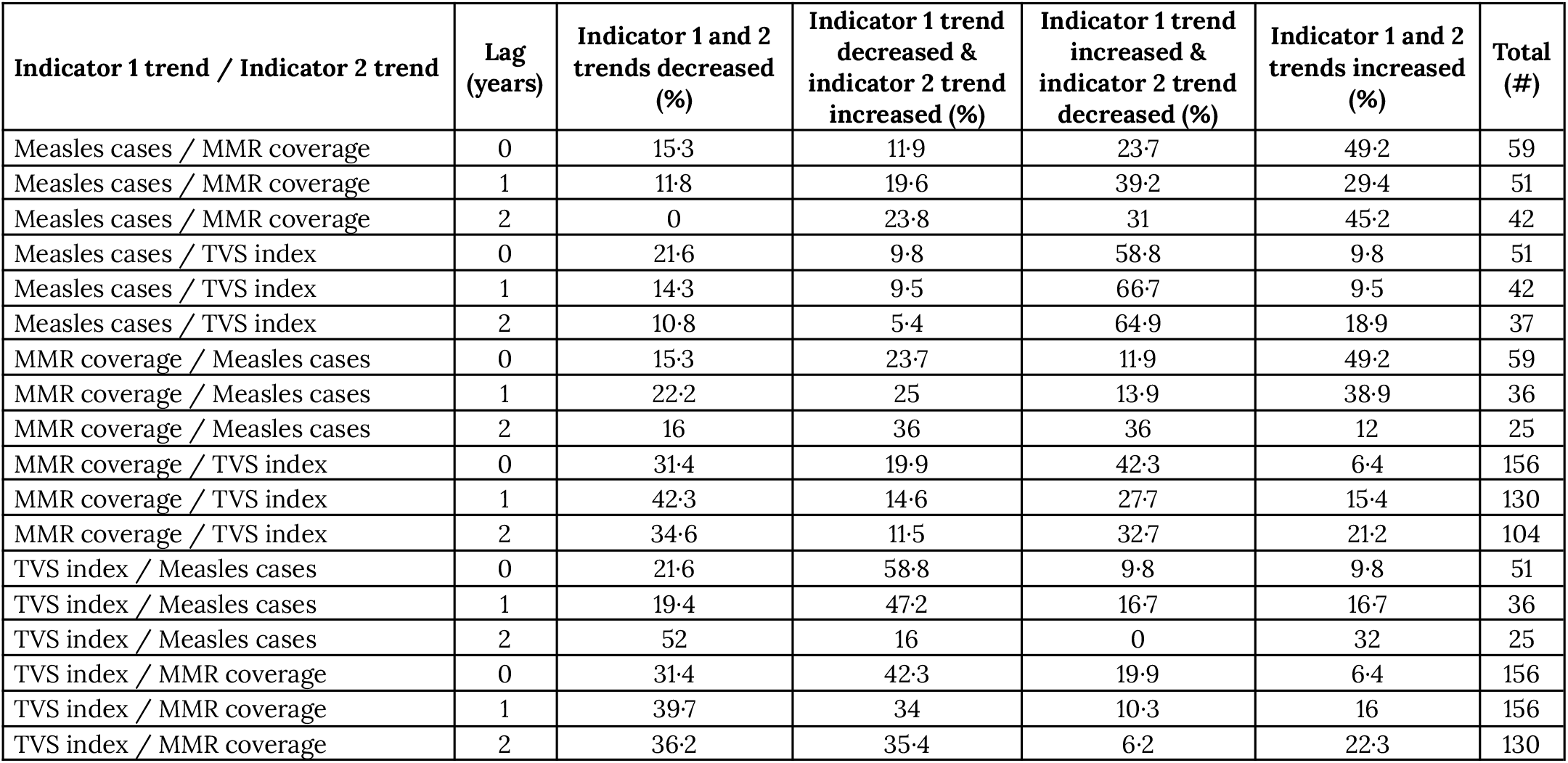
Pairwise comparison of trends of confirmed measles cases, MMR vaccination coverage of the first dose and TVS index with a lag of zero, one and two years.

| **Indicator 1 trend / Indicator 2 trend** | **Lag (years)** | **Indicator 1 and 2 trends decreased (%)** | **Indicator 1 trend decreased & indicator 2 trend increased (%)** | **Indicator 1 trend increased & indicator 2 trend decreased (%)** | **Indicator 1 and 2 trends increased (%)** | **Total (#)** |
| --- | --- | --- | --- | --- | --- | --- |
| Measles cases / MMR coverage | 0 | 15·3 | 11·9 | 23·7 | 49·2 | 59 |
| Measles cases / MMR coverage | 1 | 11·8 | 19·6 | 39·2 | 29·4 | 51 |
| Measles cases / MMR coverage | 2 | 0 | 23·8 | 31 | 45·2 | 42 |
| Measles cases / TVS index | 0 | 21·6 | 9·8 | 58·8 | 9·8 | 51 |
| Measles cases / TVS index | 1 | 14·3 | 9·5 | 66·7 | 9·5 | 42 |
| Measles cases / TVS index | 2 | 10·8 | 5·4 | 64·9 | 18·9 | 37 |
| MMR coverage / Measles cases | 0 | 15·3 | 23·7 | 11·9 | 49·2 | 59 |
| MMR coverage / Measles cases | 1 | 22·2 | 25 | 13·9 | 38·9 | 36 |
| MMR coverage / Measles cases | 2 | 16 | 36 | 36 | 12 | 25 |
| MMR coverage / TVS index | 0 | 31·4 | 19·9 | 42·3 | 6·4 | 156 |
| MMR coverage / TVS index | 1 | 42·3 | 14·6 | 27·7 | 15·4 | 130 |
| MMR coverage / TVS index | 2 | 34·6 | 11·5 | 32·7 | 21·2 | 104 |
| TVS index / Measles cases | 0 | 21·6 | 58·8 | 9·8 | 9·8 | 51 |
| TVS index / Measles cases | 1 | 19·4 | 47·2 | 16·7 | 16·7 | 36 |
| TVS index / Measles cases | 2 | 52 | 16 | 0 | 32 | 25 |
| TVS index / MMR coverage | 0 | 31·4 | 42·3 | 19·9 | 6·4 | 156 |
| TVS index / MMR coverage | 1 | 39·7 | 34 | 10·3 | 16 | 156 |
| TVS index / MMR coverage | 2 | 36·2 | 35·4 | 6·2 | 22·3 | 130 |

## 4. Discussion

This study provides an overview of the public discourse on vaccination in Brazil between 2013 and 2019 by leveraging social media data, Twitter more specifically, and large language models. The findings reveal the interrelation between public stance towards vaccination, vaccination coverage and resurgence of vaccine-preventable diseases, highlighting the usefulness of near-real-time stance for public health interventions and strategies.

The analysis of TVS index over the study period showed noticeable geographical and temporal variations in public stance towards vaccination. At national level, the overall stance was mostly neutral with fluctuations in positive and negative stance over the years. The increase of negative stance beginning in 2017 and peaking in 2019 coincides with a resurgence of measles, an eliminated vaccine-preventable disease in the country and region of the Americas. At state level, Rio de Janeiro and Rio Grande do Sul showed negative TVS indices in several years, which can be a signal of regions that may have benefited from localized public health outreach and RCCE campaigns.

The geographic findings highlight the uneven distribution of tweets across states, with Sao Paulo and Minas Gerais contributing to most of the data. These states are also some of the most populated states in Brazil. This geographic disparity in tweets also aligns with the population density and access to social media platforms, which may not allow to better understand the public discourse of vulnerable populations not represented or served by these platforms. Moreover, it underscores the necessity of having localized RCCE interventions to be able to tackle enablers and barriers for vaccination uptake specific for the population.

The temporal analysis of stance towards vaccination in the tweets during the study period showed that there were some positive peaks in the TVS index in 2018, which may reflect the positive public perception of vaccination campaigns during the 2018 yellow fever outbreak when vaccines were strongly associated with positive immediate health outcomes. On the other hand, the significant increase in negative sentiment in 2019 coincided with the measles outbreak and subsequent vaccination campaigns and loss of measles-free status, possibly driven by public concerns and the spread of incorrect information.^18^

Our findings on MMR vaccination coverage reveal critical gaps in immunisation levels across Brazilian states, especially since 2016. While all states reached the WHO-recommended 95% coverage for the first dose in 2014, it was not sustained in subsequent years. In 2018 and 2019, few states met the target, coinciding with the measles outbreaks in those years. These trends align with the decline in TVS index, suggesting a possible relation between decreased public stance towards vaccination and declining vaccination coverage.

The geographic distribution of confirmed cases of measles further illustrates this relation. States with lower TVS indices and reduced vaccination coverage, such as Amazonas, Pará and Roraima, experienced the highest number of confirmed measles cases during the outbreak.

However, when analysing the pairwise trends of TVS index, confirmed measles cases and MMR vaccination coverage of the first dose there was no clear pattern of any pair of trends, except for increased confirmed measles cases followed by decreased TVS trends in the same year or one or two years after. This reiterates the importance of considering the vaccination acceptance an outcome of multifactor determinants, including non-health related determinants as indicated in previous studies.^19^ This also aligns with the predominant discourse found in this study in the tweets with a negative stance towards vaccination in which there were mentions on non-healthcare factor such as the management of epidemics and vaccination by the government.

Brazil losing the measles-free status in 2019, following the 2018 outbreak and decrease of stance towards vaccination and decrease in MMR vaccination coverage highlights the need for sustained and continuous public health efforts, integrating near-real-time monitoring of public discourse and stance towards vaccination, and targeted RCCE interventions to ensure sufficient vaccination coverage even when the disease is considered eliminated.

The main limitations of this study are the use of one online platform with uneven distribution of tweets per state. We have addressed this by doing a specific analysis per state avoiding using the results from national data as equally to those from the different Brazilian states. Although we have covered only one online social media platform, it settles the ground bases for expanding to other sources easily, online or offline.

While this study focused on Twitter data, Brazil, and a specific timeframe, this approach can be easily applied to other data sources, contexts, and time periods, providing a more holistic and inclusive overview of the public discourse towards vaccination. Likewise, it could help in assessing how epidemics or pandemics such as the COVID-19 pandemic reshape the public discourse around vaccination.

Following results from our previous research on the capacity of GPT-4 to assess vaccination-related tweets and its stable performance using different prompts, we chose the model to extract the stance towards vaccination from the tweets’ text.^15^ Considering that some models are sensitive to the prompt and can have different performance results depending on the data and objective, the usage of a previously evaluated LLM on the same use case (stance towards vaccination using tweets) which were not impacted by the prompt used allowed us to choose the best combination of model and prompt for our study. This was a key aspect since there is currently no publicly available benchmark for specific use cases in public health. The integration of LLMs into public health workflows allows processing vast amounts of data in near-real-time, providing timely and actionable insights for identifying behavioural enablers and barriers to vaccination and adapt public health communication strategies accordingly.

In conclusion, this study has shown the potential of a near-real-time public stance towards vaccination to inform RCCE strategies, more specifically related to vaccination. By understanding community perspectives and needs, public health authorities can develop responsive and adaptive public health approaches to provide accurate information and support informed vaccination decisions. Future research should combine online and offline data sources and assess how the public health stance is affected by epidemics or pandemics.

## Contributors

LE was responsible for the data curation, formal analysis, methodology, visualization, writing original draft of the manuscript, and reviewing and editing the final version of the manuscript. MS was responsible for the conceptualization, funding acquisition, methodology, supervision, and reviewing and editing the final version of the manuscript.

## Data Availability

The data (list of tweets identifiers and summarised anonymised datasets) and R and Python code used can be found in the online repository at https://github.com/digitalepidemiologylab/brazil-vaccine-uptake.

## Funding

This work was supported by Fondation Botnar and the European Union’s Horizon H2020 grant VEO (874735). The funders had no role in the design or execution of this study and will have no role in the analyses, interpretation of the data, or decision to submit results.

## Declaration of interests

The authors declare no conflict of interest.

## Acknowledgements

We would like to acknowledge Dr. Ana Rivière Cinnamond from the Pan American Health Organization (PAHO) for the discussions which contributed to the selection of the study location and keywords for data collection.

## Notes

### Competing Interest Statement

The authors have declared no competing interest.

### Funding Statement

This work was supported by Fondation Botnar and the European Union Horizon H2020 grant VEO (874735). The funders had no role in the design or execution of this study and will have no role in the analyses, interpretation of the data, or decision to submit results.

### Author Declarations

This work has been approved by the EPFL Human Research Ethics Committee (HREC No. 012.2018 / 05.03.2018). Crowdbreaks anonymises the text of all tweets changing the mention or reference to usernames to @username and the URLs to url, and complies with the Twitter terms of service.

